# Oral health professionals and child abuse and neglect: a scoping review protocol

**DOI:** 10.1101/2022.03.18.22272542

**Authors:** Heuiwon Han, Zac Morse, Jane Koziol-McLain, Amanda B Lees

**Affiliations:** Department of Oral Health, School of Clinical Sciences, Auckland University of Technology, 90 Akoranga Drive, Northcote, Auckland, New Zealand; Centre for Interdisciplinary Trauma Research, Auckland University of Technology, Auckland, Aotearoa New Zealand; School of Public Health and Interdisciplinary Studies, Auckland University of Technology, Auckland, Aotearoa New Zealand

## Abstract

**Introduction:** Child abuse and neglect can seriously impair social development, learning, physical, and psychological health. Oral health practitioners can recognize the orofacial manifestations of maltreatment in a dental setting, provide necessary support, and refer concern to child protection agencies to prevent further harm. There is a need to understand the responsiveness of oral health practitioners to a child protection concern. This protocol outlines the systematic processes to map evidence in the field of child protection in oral health.

**Methods and analysis:** This review will be guided by the PRISMA-ScR and the JBI guidelines. Primary and secondary studies, reviews, guidelines, and editorial articles written in English will be included in the review. A three-step search will be implemented. An initial search is conducted to finalised the search terms and eligbility criteria followed by a second search involving sensitive searches of databases (Scopus, MEDLINE, CINAHL, Dentistry & Oral Science Source, and Cochrane Library). A third search will involve grey literature searching for unpublished materials. Government and international health organization websites will be searched during the third search. Eligible studies will be screened based on the participants (oral health practitioners), the concept (child protection) and the context (dental setting) and data extracted by two reviewers. Outcomes from the data extraction chart will be synthesized through exploratory narratives.

**Ethics and dissemination:** This study involves neither human participants nor unpublished secondary data. As such, ethics approval is not required. Findings will be disseminated through publication in a peer-reviewed journals, professional networks, and conferences.

**Strengths and limitations of this study:**

- This study will systemically identify literature from a broad range of databases and map evidence in the field of child protection in oral health.
- The review will take a rigorous approach adhering to the PRISMA-ScR and and the JBI guidelines.
- Multiple reviewers will conduct screening and data extraction independently to facilitate reliability.
- A limitation is that only sources in English will be reviewed.

## INTRODUCTION

Children and young people’s right to safety and health are enshrined in the United Nations Convention on the Rights of the Child 1993. Children and adolescents’ age and developmental status, however, means that adults are responsible for protecting against abuse. Children’s rights are vulnerable to being violated when adequate care is not provided. The World Health Organization defines child abuse and neglect (CAN) as:

“All forms of physical and/or emotional ill-treatment, sexual abuse, neglect or negligent treatment or commercial or other exploitation, resulting in actual or potential harm to the child’s health, survival, development, or dignity in the context of a relationship of responsibility, trust, or power.”^1(p15)^

The definition includes any act or failure to act that can endanger children’s optimum health, survival, or development.^1^ Four types of CAN are commonly recognized: physical abuse, sexual abuse, emotional abuse, and neglect. The United Nations Office on Drugs and Crime (UNODC)^2^ reported that an estimated total of 205,153 children aged 0 to 15 years lost their lives worldwide as a result of homicide between 2008 and 2017. Homicidal acts were mainly perpetrated by family members or people who were close with the victims. The UNODC estimated that up to one billion children aged 2 to 17 years experienced child abuse and neglect in 2017.^2^

CAN experiences can seriously impair social development, learning, short- and long-term physical and psychological health.^1 3-6^ An analytic review of international literature conducted by Leeb *et al*. ^4^ reported that 50% to 90% of abusive or neglected cases do not come to the attention of child protection authorities which lead to potential detrimental impacts. Consequences include health outcomes such as fractures, lacerations, and central nervous system injuries, as well as emotional developmental impairments, sexual dysfunction, and depression.^3 4 6-9^ A systematic review and meta-analysis by Norman *et al* ^10^ indicated significantly increased risks of developing depressive mental disorders when physically abused (OR = 1.45, 95% CI 1.16-2.04) and emotionally abused (OR = 3.06, 95% CI 2.43-3.85). Moreover, the victim of physical abuse (OR 2.58, 95% CI 1.17-5.70) and emotional abuse (OR = 2.56, 95% CI 1.41-4.65) was more likely to experience an eating disorder as a long-term health consequence.^10^

In the context to protect children from CAN, oral health practitioners have an important role in safeguarding children.^4 7 11 12^ Oral health practitioners are in a strategic position to recognize the orofacial manifestations of CAN, support children and their families and refer potential victims to child protection agencies if necessary.^13^ The systematic review carried out by Sarkar *et al*.^5^ of the patterns of orofacial injuries concluded that different facial and intraoral indications, including abrasions, contusions, and lacerations, are commonly presented in child physical abuse. Furthermore, multiple oral lesions and healing spots may indicate exposures to physical or sexual abuse.^14^ Non-accidental traumatic injuries around the head and neck areas can be detected during dental visits. Furthermore, behavioural and mental health manifestations of CAN, including excessive defensive or aggressive behaviour and excessive fear of caregivers, can be assessed during routine dental examinations.^14 15^ As defined by the American Academy of Pediatric Dentistry^16(p16)^, dental neglect is the “willful failure of a parent or guardian to seek and follow through with treatment necessary to ensure a level of oral health essential for adequate function and freedom from pain and infection”. Untreated oral conditions can lead to impaired learning, growth, and nutrition.^17^ Oral health providers are well-placed to assess caregiver barriers to care and their knowledge and awareness of the needs for oral health attention. A child’s social, emotional, and medical conditions should be carefully considered when determining dental neglect.^17^ This requires knowledge and emphatic attitudes of oral health practitioners. Early intervention and prevention approaches can be designed and implemented in clinical practices in ways that suit the community and the profession.

Even though oral health practitioners can play a critical role in child protection, under-reporting of suspected cases is evident globally. For example, survey research from Denmark (n = 1145) of Danish dentists and hygienists indicated that 38.3% of the respondents suspected CAN during their careers; however, only 33.9% of those who suspected CAN reported their concern to child protection agencies for further investigation.^18^ Similar findings from other surveys have been reported from various parts of the world. A study from Turkey (n = 1020) revealed 17.1% of the respondents (dentists, specialists, and dental students) suspected CAN in the last five years, while only 1% reported^19^; a Saudi Arabian study (n = 122) revealed 59% with suspected cases and with approximately 10% reported cases in the last five years^20^; United Kingdom study (n = 105) revealed 15% with suspected cases and with 6% reported cases in the last six months.^21^ Rates of detecting suspected cases vary among countries, yet under-reporting of CAN by oral health practitioners is evident worldwide.

A systematic review was conducted by Rodrigues and co-authors^22^, which confirmed the under-reporting of suspected CAN by oral health practitioners and insufficient knowledge to detect CAN cases. A scoping review by Bradbury-Jones and co-authors^23^ confirmed the relationship between CAN and poor oral health and head and neck trauma as common CAN indicators. They commented on the need to improve the knowledge and confidence of oral health practitioners to respond to CAN and the need for interdisciplinary practice and continuous learning related to the topic.^23^ Yet, authors reported that discussion papers, editorials or policy documents were excluded and recommended further explorations of the topic.^23^

The authors believe a broad scoping of the existing literature would help to further explore child protection in dental settings. This study will involve mapping how oral health practitioners are responding to child abuse and neglect concerns in the dental setting. As there is limited understanding of the evidence in child abuse and neglect in dental settings, the proposed review will address this gap, enabling the identification of areas where more primary research is needed.

## METHODS AND ANALYSIS

A systematic scoping review maps the literature and provides an overview of the evidence, concepts, or studies in a particular field.^24 25^ The results of a scoping review can provide indications for further research and inform the development of research endeavours.^24 26^ Scoping newly emerging evidence about oral health practitioners towards CAN will provide a greater overview of the topic and guide future areas to focus. A preliminary search of MEDLINE, CINAHL, Cochrane Database of Systematic Reviews, Open Science Framework Database, JBI Evidence Synthesis, JBI Systematic Review register, and PROSPERO was conducted to check current or ongoing systemic reviews or scoping reviews of the topic. The scoping review will adopt the Joanna Briggs Institute (JBI) guideline by Peters and co-authors^27^, which was influenced by Arksey and O’Malley^28^ and Levac and co-authors.^29^ The guideline comprises the six stages which include (1) identifying the research question, (2) identifying relevant studies, (3) select studies, (4) chart the data, (5) collate, summarise and report the results, and (6) consultation ^27^. The stage six (consultation) will be considered after reporting the results. The Preferred Reporting Items for Systemic Reviews and Meta-Analyses extension for Scoping Reviews (PRISMA-ScR) will be referred to ensure rigour and replicability of the review. ^30^

### Stage 1: identifying the research questions

The primary question of this scoping review is to examine and map child abuse and neglect and child protection in dental settings. Through consultation with the research team, the following sub-questions were identified:

1. What is the role of oral health practitioners in child protection?
2. What is the current knowledge and attitudes oral health practitioners towards CAN?

As conducting a scoping review is iterative, the research team will become increasingly familiar with the topic. Therefore, the research question and sub-questions may be revised and amended. Factors including barriers, facilitators, interventions, and needs of the practitioners will be considered for the review.

### Stage 2: identifying relevant studies

After consultation between the research team and the Auckland University of Technology librarian, the initial search strategy was developed from the preliminary search outcome (Appendix I). The search strategy was underpinned by key inclusion criteria (**Table 1**). Eligibility criteria were categorized by the Population/Concept/Context (PCC) mnemonic with types of evidence sources^27^.

**Table 1.** Review eligibility criteria based on study population, concept, context, and type of evidence

| Element | Inclusion criteria |
| --- | --- |
| Population | Oral health practitioners who are registered with national regulatory bodies (dentists, dental specialists, oral health therapists, dental therapists, dental hygienists, and orthodontic auxiliaries)<br>Undergraduate and postgraduate students for dental-related programmes<br>Other parties that are involved in child oral health care |
| Concept | Child abuse and neglect<br>Child protection |
| Context | All international dental-related settings, including private and public dental services<br>Dental services provided in community settings such as schools |
| Types of evidence | Primary and secondary research studies<br>Systematic reviews and metanalyses<br>Editorial articles<br>Government policies and guidelines<br>International organization guidelines<br>English, full-text |

The search strategy involves three steps.^27^ The first initial limited search involves searching two online databases (Medline and CINAHL) to identify relevant topics, followed by analyzing the text wording in the title, abstract, and index terms of identified literature. The findings from the initial search will inform the search terms and eligibility criteria. After the first initial search, two independent reviewers will pilot the selection process using eligibility criteria with one database (MEDLINE) to ensure consistency and validity of the search. The first 25 sources will be screened by their titles and abstracts and labelled as ‘included’, ‘excluded’ or ‘uncertain’. If differences arise, eligibility criteria and selection process will be reviewed and consulted by all researcher. The second search involves searching all databases (Scopus, MEDLINE, CINAHL, Dentistry & Oral Science Source, and Cochrane Library). The search will include any relevant sources up to the end of 2021. Then, a follow-up search will be conducted at a later date to identify any source published in 2022. The third search will be conducted to identify any additional studies by examining the reference lists of the sources that have been selected for data extraction. The researchers may contact the authors of primary studies or reviews for further information if this is relevant. After the third search, grey literature searching will be done to identify unpublished materials relevant to the topic. Government websites and international health organization websites will be searched. The citation records from databases will be exported into EndNote X9, and duplicates will be removed. Data will then be imported to Covidence to manage the screening and extraction processes.

### Stage 3: study selection

After completing second and third searches involving all databases, two reviewers will independently review titles and abstract against included and excluded criteria. Once all titles and abstracts are screened, two reviewers will independently assess the first ten full-text articles for eligibility. If differences arise, eligibility criteria and the selection process will be reviewed and there will be consultation with all researchers. Once agreement is achieved, all the full-text articles will be independently reviewed by two reviewers for inclusion and exclusion. Any uncertainty will be discussed with the team. Reasons for exclusion of any research during the full-text screening will be recorded, which will be provided in an appendix of the final review report.

### Stage 4: data extraction

The JBI guideline^27^ will inform data extraction and charting which will be managed in Covidence Data charting (Appendix II) will include authors, year of publication, country of origin, aims and purpose, study population, sample size, methodology and methods, intervention (details, durations, and outcomes), key findings related to the review questions, and future recommendations. The content of the data charting form will evolve as more understanding is gained throughout the process. Therefore, the table will be reviewed after the study selection.

To ensure consistency and accuracy, the data charting form will be trialled on three sources by two reviewers. Any difference will be discussed as a team; then, a single form will be finalized. Once agreement is achieved, one reviewer will complete data extraction and charting for other sources, and the second reviewer will review the form afterwards. Any disagreement will be discussed as a team and revise the data charting form.

### Stage 5: collating, summarising and reporting the result

Following data extraction, a descriptive analysis will be carried out to map the finding. A method flow chart will be produced according to the PRISMA-ScR guideline.^30^ Findings will be summarised and presented in a tabulated form configured in such a way as to address the objectives of the review.^27^ Data will be accompanied by an exploratory narrative focusing on the role of oral health practitioners in child protection, responsiveness, experiences, and barriers and facilitators. The reviewers will discuss the implications of the findings on further research, practice, and policy.^29^

## Data Availability

All data produced in the present work are contained in the manuscript.

## ETHICS AND DISSEMINATION

This study involves neither human participants nor unpublished secondary data. As such, ethical approval is not required. Findings of the scoping review will be disseminated through publication in a peer-reviewed journal, professional networks, and conferences.

## Acknowledgements

The authors thank Andrew South, Liaison Librarian for Health at the Auckland University of Technology, for their assistance in developing the search strategy for this study.

## Appendix I: Initial search strategy

|  | Search terms | CINAHL, Dentistry & Oral<br>Science Source, MEDLINE |
| --- | --- | --- |
| 1. Population | dentis* or dental professionals or<br>dental practitioners or oral health<br>practitioners AND | 797,125 |
| 2. Concept | children or adolescents or youth or<br>child or teenager or teens or young<br>people or kids or paediatric or<br>pediatric AND | 5,464,088 |
| 3. Concept | abus* or neglect* or maltreat* or<br>safeguard* AND | 466,990 |
| 1 & 2 & 3 = 2,547 |  |  |

## Appendix II: data charting form

|  |
| --- |
| **Basic information** |
| Title |
| Citation |
| Authors |
| Year of publication |
| Country of origin |
| Industry funding (Y/N/Not specified) – if Y, role of funders in the study |
| Aims and objectives |
| Study design (RCT, cohort, review, etc) |
| Methodology |
| Study population (if applicable) |
| Sample size (if applicable) |
| Intervention (details and duration) (if applicable) |
| Data collection method |
| Data analysis method |
| **Key findings** |
| Role of oral health practitioners in child protection |
| Reporting and detecting of CAN |
| Knowledge and attitudes towards CAN |
| Barriers and facilitators |
| Needs of oral health practitioners to improve their responsiveness |
| Any interdisciplinary collaboration |
| Any effort to reduce inequities in child protection |
| Other findings |
| Recommendations (if applicable) |
| Strengths and limitations |

